# Evaluation of Seropositivity after Standard Doses of Vaccination against SARS-CoV-2 in Early Breast Cancer Patients Receiving Adjuvant Treatment

**DOI:** 10.1101/2022.07.23.22277888

**Authors:** Jinyong Kim, Euijin Chang, Song Yi Park, Dae-Won Lee, Chang Kyung Kang, Pyoeng Gyun Choe, Nam Joong Kim, Myoung-don Oh, Wan Beom Park, Kyung-Hun Lee, Seock-Ah Im

**Author notes:** Co-first authors (Jinyong Kim, Euijin Chang). **Correspondence to** Kyung-Hun Lee, MD, PhD, Department of Internal Medicine, Seoul National University Hospital, 101, Daehak-ro, Jongro-gu, Seoul 03080, Republic of Korea, Wan Beom Park, MD, PhD, Department of Internal Medicine, Seoul National University Hospital, 101, Daehak-ro, Jongro-gu, Seoul 03080, Republic of Korea. **Author Contributions** K.L. and W.P. conceptualized the study and set methodology. J.K. and E.C. performed data collection, formal analysis and interpretation. All authors performed data curation. K.L. and W.P. supervised the study. J.K. and E.C. wrote original manuscript. All authors reviewed and provide final approval of the manuscript.

## Abstract

**Background:** Coronavirus disease 2019 (COVID-19) pandemic affected millions of individuals and patients with cancer are known to be more susceptible. Vaccines against SARS-CoV-2 have been developed and used for patients with cancer, but scarce data is available on their efficacy in patients under active anti-cancer therapies.

**Materials and Methods:** In this study, we semi-quantitatively measured the titers of the immunoglobulin G against the anti-spike protein subunit 1 of SARS-CoV-2 after vaccination in early breast cancer patients with concurrent chemotherapy, endocrinal or targeted non-cytotoxic treatments, and no treatments.

**Results:** Standard doses of COVID-19 vaccines provided sufficient immune responses in patients with early breast cancer, regardless of the type of anticancer therapies. However, the post-vaccination serum anti-spike antibody titers were significantly lower in the patients under cytotoxic chemotherapy.

**Conclusion:** Our study emphasizes the importance of the personalized risk stratification and consideration for booster doses in more vulnerable populations.

**Implications for Practice:** In this article, we present rare data on the homogeneous population of patients with early breast cancer under active anti-cancer treatments. The patients showed adequate serologic responses against SARS-CoV-2 virus after standard doses of vaccination without serious adverse events in concurrence with active adjuvant anti-cancer therapy. The patients receiving concurrent cytotoxic chemotherapy, however, have significantly lower serum anti-spike antibody titers than those under non-cytotoxic anti-cancer treatments or without treatments. Selection of the COVID-19 vaccines should be based on personalized risk stratification considering the use of concurrent cytotoxic chemotherapy and the patient’s medical circumstances. Booster doses of vaccination could be considered for the vulnerable population, such as elderly with comorbidities and the single standard dose of vaccination.

## Introduction

Coronavirus disease 2019 (COVID-19) pandemic had affected over the world with 308 million confirmed cases and 5.49 million deaths as of January 12, 2022 ^1^. Standard doses of authorized vaccines against severe acute respiratory syndrome coronavirus 2 (SARS-CoV-2), which included BNT162b2 vaccine (Pfizer–BioNTech (PZ)), mRNA-1273 vaccine (Moderna), Ad26.COV2.S vaccine (Janssen)^2^, and AZD1222 vaccine (Oxford-AstraZeneca (AZ)), provided comparable efficacy in preventing symptomatic COVID-19 with the acceptable safety profile^3-6^.

Patients with cancer have an increased mortality rate due to COVID-19, and are considered as a prioritized group in need of vaccination by most authorized global guidelines ^7-12^. Nonetheless, patients with cancer under systemic treatments were largely excluded from the previous clinical trials conducted to assess the efficacy of the COVID-19 vaccines. Although recent studies reported that the standard dose of vaccine induced adequate responses in patients with cancers, data on each specific solid cancer needs further investigation ^13,14^.

Breast cancer is the leading cause of the cancer deaths in female ^15^. Especially, the early breast cancer confined to the breast and regional lymph nodes accounts for majority of the total cases ^16,17^. Treatments of early breast cancer are usually composed of various modalities including surgery, radiotherapy, hormonal therapy, molecular targeted therapy, and chemotherapy with corticosteroid prophylaxis ^16^. Thus, the effect of COVID-19 vaccination should be reevaluated among the patients with early breast cancer who are currently under different anti-cancer treatments.

This study is a prospective study evaluating the serologic responses from standard doses of COVID-19 vaccines in the patients with early breast cancer, by comparing the antibody responses in the patients on cytotoxic chemotherapy with those in the patients on the other anti-cancer treatments. We also investigated whether the adjunct corticosteroid affected the immunologic responses to vaccines in the patients with early breast cancer.

## Materials and methods

### Study design and subjects

This is a prospective, longitudinal cohort study that commenced recruitment in April 2021 at Seoul National University Hospital. Early breast cancer patients who are 20 years or older with the past or current history of systemic anti-cancer treatments for breast cancer, including cytotoxic chemotherapy, molecule-targeted therapy, and endocrinal therapy were enrolled. Early breast cancer was defined as the disease confined to the breast with or without regional lymph node involvement, and without distant metastatic disease. The patients who were currently under chemotherapy within 28 days of vaccination were included in the cytotoxic chemotherapy group; patients under targeted therapy and endocrinal therapy were in non-cytotoxic anti-cancer treatment group; patients who did not receive any anti-cancer treatment within 28 days of vaccination were in control group. The patients received standard doses of vaccines, which were either two doses of PZ, Moderna, AZ, or a single dose of Janssen. The patients who had active infectious diseases or received immune-suppressants or organ transplant were excluded.

The chemotherapy regimen included anthracyclines, cyclophosphamide, docetaxel, paclitaxel, carboplatin, and capecitabine. Molecular target agents included trastuzumab and pertuzumab. Hormonal therapies were chosen among tamoxifen, letrozole, and anastrozole depending on the patient’s menopausal status and clinical conditions. The association between corticosteroid use and vaccination was also assessed in the patients who used corticosteroid over 10mg of prednisolone or equivalent dose within 14 days of vaccination ^14^.

Our primary endpoint was the immunogenicity of the COVID-19 vaccine in the patients with early breast cancer under active systemic anti-cancer treatment, defined by the proportion of patients who achieved positive anti-spike (anti-S1) antibody titer after vaccination. Secondary outcomes included the association of the prior use of corticosteroids to immunogenicity of vaccines. Clinical data and samples were collected before the vaccination, between 21 to 28 days after the first and the second dose of vaccination, respectively. The study was approved by the Institutional Review Board (IRB No. 2103-121-1206). Written informed consent was obtained from all participants prior to vaccination. All the data from the patients were anonymized and de-identified prior to analysis.

### S1-reactive IgG ELISA

We evaluated the antibody responses at baseline and at 3 weeks after each vaccination. We semi-quantitatively measured the concentration of the immunoglobulin G against the anti-S1 of severe acute respiratory syndrome coronavirus 2 (SARS-CoV-2) using Enzyme-linked immunosorbent assay (anti-S1 IgG ELISA) (Euroimmun, Lübeck, Germany) according to the manufacturer’s instructions. The immunoassay was granted Emergency Use Authorization by the US Food and Drug Administration. The cut-off index (COI) of optical density ratio (OD ratio) greater than or equal to 1.1 was considered seropositive; COI lower than 0.8 was considered negative; and the rest was considered borderline according to the manufacturer’s instructions. Borderline results were considered as negative for analysis.

### Statistical analyses

Categorical variables were summarized with the frequencies in number and rates in percentages. Continuous variables were represented with the median values and ranges. Differences were assessed using Mann-Whitney test for continuous variables and Pearson’s χ2 or Fisher’s exact test for categorical variables. P-values of <0.05 were considered statistically significant. The association between treatment with cytotoxic chemotherapy and the anti-S1 IgG OD ratio in post-vaccination serum was evaluated using a multivariate linear regression model. The anti-S1 IgG OD ratio was set as the dependent variable and the treatment with cytotoxic chemotherapy was set as the independent variable. Additional covariates were age and type of vaccines. The association between treatment with cytotoxic chemotherapy and the seropositivity based on OD ratio was evaluated using a logistic regression model. Data and statistical analyses were performed in R version 4.1.2 and RStudio version 2021.09.1.

## Results

### Patient characteristics

We recruited 112 patients with early breast cancer who received adjuvant cytotoxic chemotherapy and at least one dose of COVID-19 vaccine. Twelve patients withdrew from the study, and another twelve samples were not available for the analysis at the time of data collection. One case was excluded because of the high baseline anti-S1 antibody level, suggestive of COVID-19 infection prior to vaccination (Figure 1). Of all, 80.5% (70 of 87) received PZ; 10.3% (9 of 87) received Moderna; 3.4% (3 of 87) received Janssen; 5.7% (5 of 87) received AZ. Except for the three patients who received single dose of Janssen, 84 patients received two doses of vaccines without cross-injection. The median interval between the first and second vaccination was 42 days (range, 21-77). The median age of patients was 52 years (range 35-69) and all were female. Within 28 days of vaccination, 36.8% (32 of 87) underwent cytotoxic chemotherapy and 9.2% (8 of 87) had combination with targeted therapy. Patients who received non-cytotoxic anti-cancer therapy were 42.5% (37 of 87). Only one patient was under molecular targeted therapy, and other 36 patients were under endocrinal therapy. The patients who had no anti-cancer treatment within 28 days of vaccination were 20.7% (18 of 87). No serious adverse events were reported. The patient characteristics are shown in Table1.

**Figure 1.**
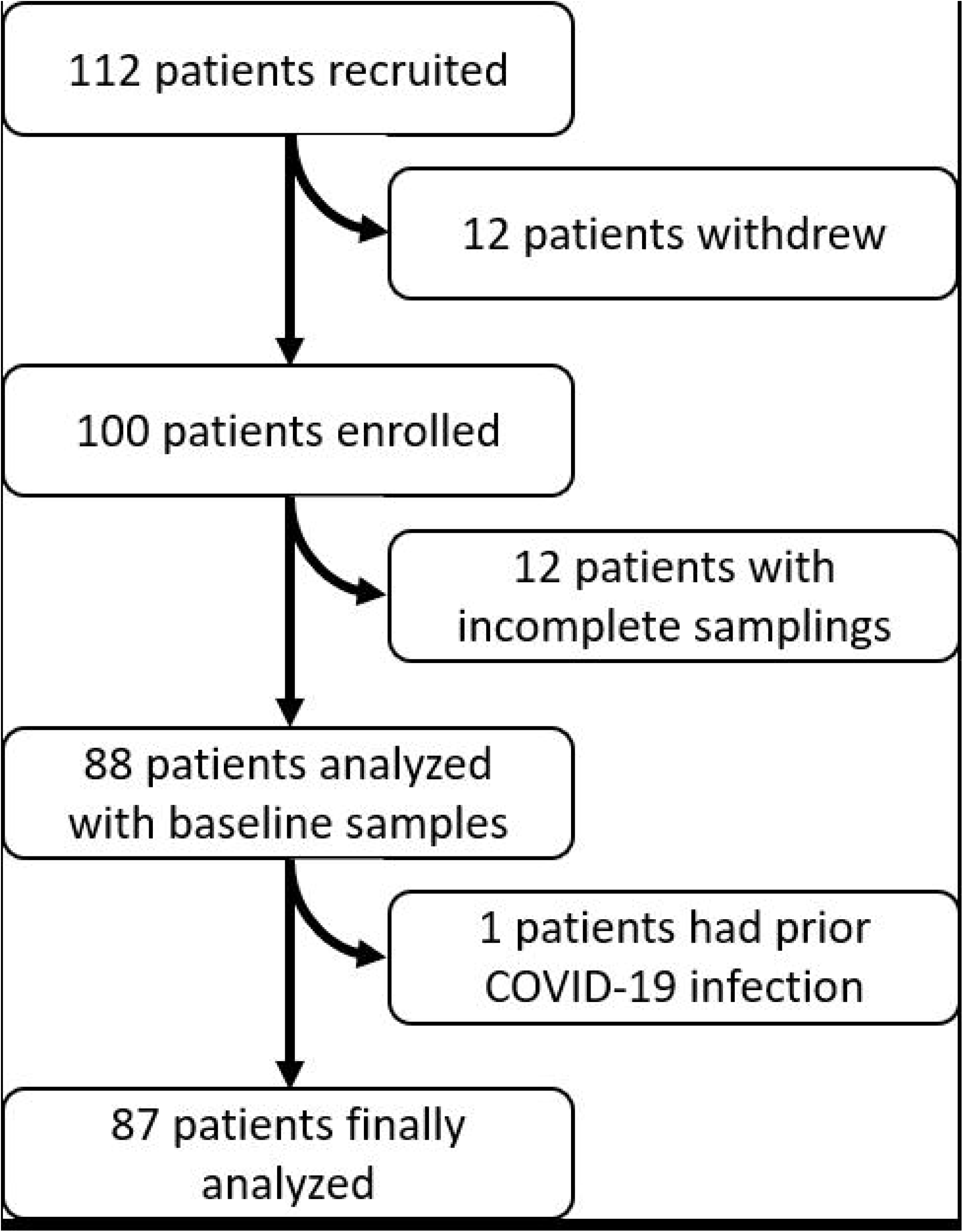
Patient enrollment: Flowchart showing the patient enrollment and selection

### Vaccine immunogenicity with active chemotherapy

Overall, 96.6% (84 of 87) were seropositive after completion of vaccinations. After first dose, 70.1% (61 of 87) were seropositive, and additional 25.3% (22 of 87) gained seropositivity after second dose (Figure 2, Table S1). Three patients failed seroconversion; two were in borderline range and one was seronegative. Two patients in borderline range received two doses of PZ vaccine during adjuvant doxorubicin and cyclophosphamide chemotherapy. The only seronegative patient received one dose of Janssen while on letrozole (Table S2).

**Figure 2.**
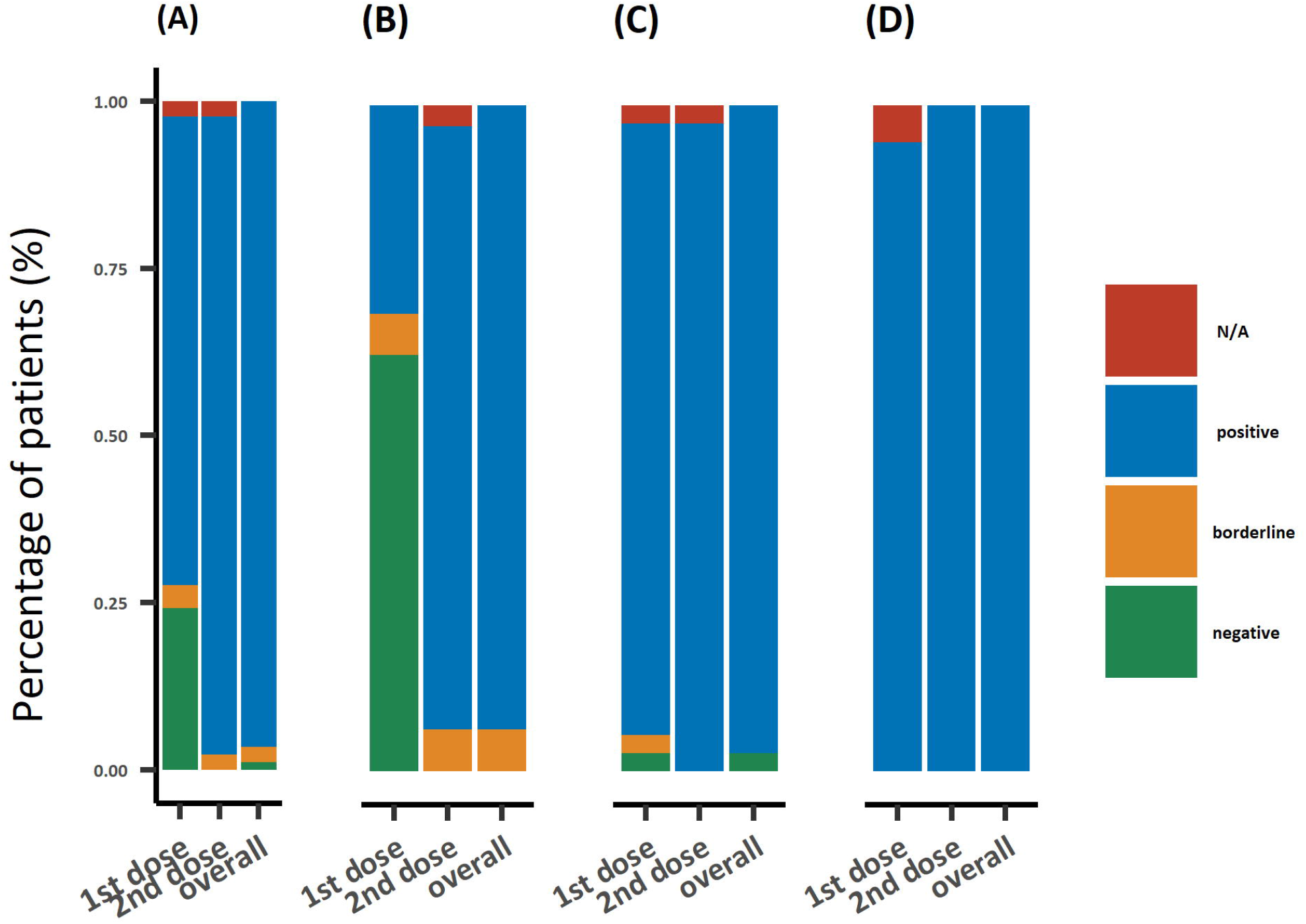
Proportion of seroconversion in patients: Seroprevalence proportions in (A) all early breast cancer patients, (B) with cytotoxic chemotherapy treatments, (C) with non-cytotoxic treatments, and (D) without anti-cancer treatment.

Only 31.2% (10 of 32) in the chemotherapy group, compared with 91.9% (34 of 37) in the non-cytotoxic treatment group and 94.4% (17 of 18) in the control group, achieved the adequate immunoglobulin level after the first dose. However, the majority (93.8%, 30 of 32) of the patients in chemotherapy group achieved seropositivity after the second dose. In the non-cytotoxic treatment group and control group, all patients who received two doses of vaccination were seropositive. There was no significant difference in seroconversion rate after the completion of vaccination among the cytotoxic chemotherapy group, non-cytotoxic treatment group, and control group (93.8% vs. 97.3% vs. 100%, p=0.234). No significant association was observed in seropositivity regarding age, types of vaccines, and treatments with cytotoxic chemotherapy (Table S3).

Anti-S1 antibody levels increased gradually with each vaccination in all patients, with more than 50 folds after the first dose and 137 folds after the second dose (Figure S1). Both mRNA or adenovirus vector vaccines showed significant increase in antibody titers (Figure S2), and no significant difference was observed in the anti-S1 antibody titer at each time point.

When the antibody titers in the chemotherapy group, the non-cytotoxic treatment group, and the control group were compared, the antibody titers also significantly and gradually increased in all groups after the first and second vaccinations. Despite similar baseline levels, however, significantly lower magnitudes of antibody titers was observed in patients who had concurrent cytotoxic chemotherapy, compared to those in the non-cytotoxic treatment group or those in the control group after first vaccination (cytotoxic chemotherapy vs. non-cytotoxic treatment vs. control, 10.7 vs. 84.2 vs. 69.8-fold increase respectively, p<0.001) and second vaccination (95.4 vs. 142.1 vs. 132.4-fold increase respectively, p<0.001 for cytotoxic vs. non-cytotoxic and p=0.005 for cytotoxic vs. control) (Figure 3, Table 2). In the linear regression and multiple regression analyses adjusted for age, the treatment with cytotoxic chemotherapy remained as a significant factor for predicting low anti-S1 antibody titer (p<0.001). No significant association was observed between the antibody titers with types of vaccines (Table S4, S5).

**Figure 3.**
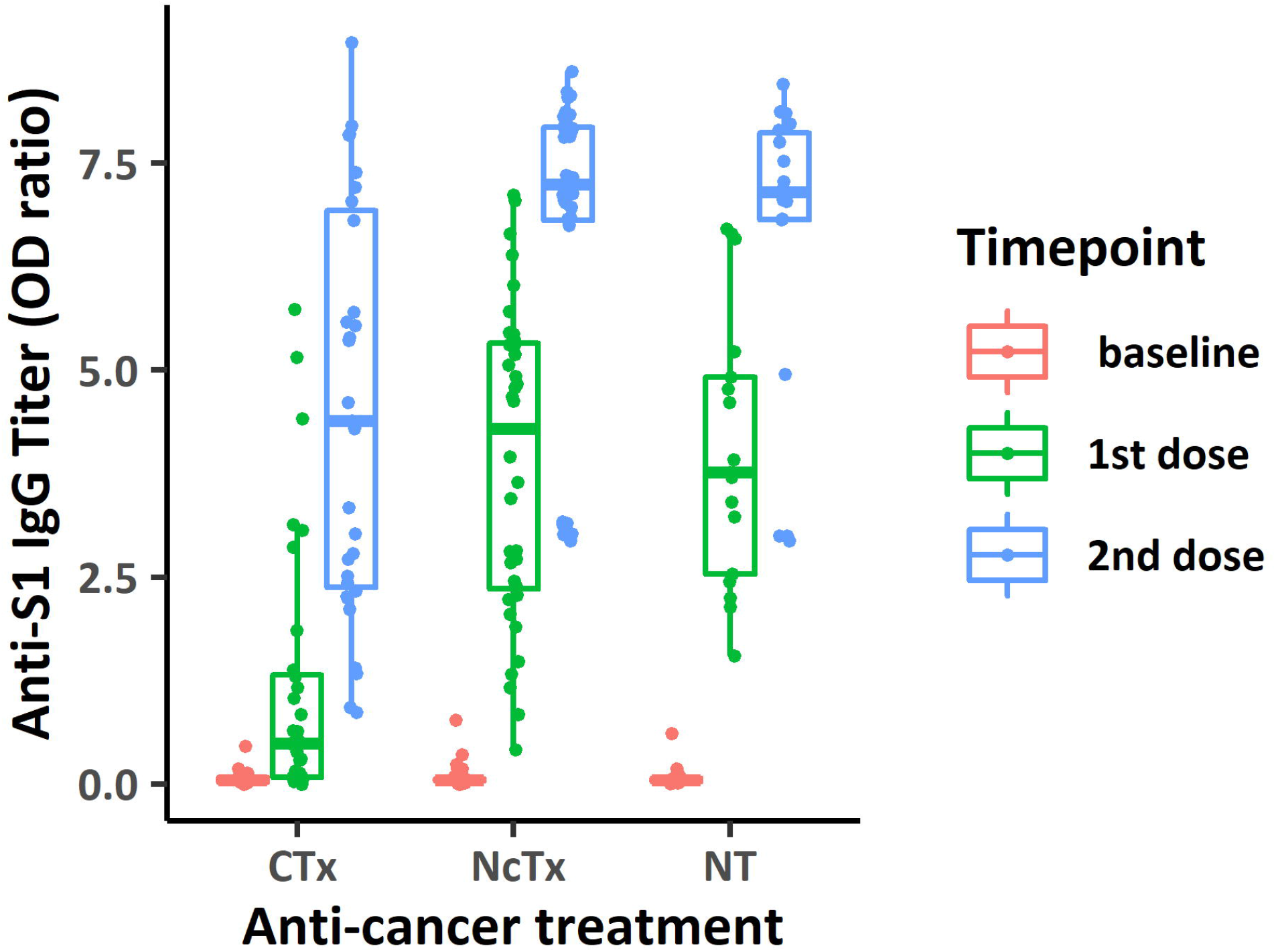
Antibody titers with Anti-Cancer treatments: SARS-CoV-2 anti-S IgG antibody titers after each dose of vaccinations in early breast cancer patients with cytotoxic chemotherapy treatment, with non-cytotoxic treatment, and without anti-cancer treatment

**Table 1.**
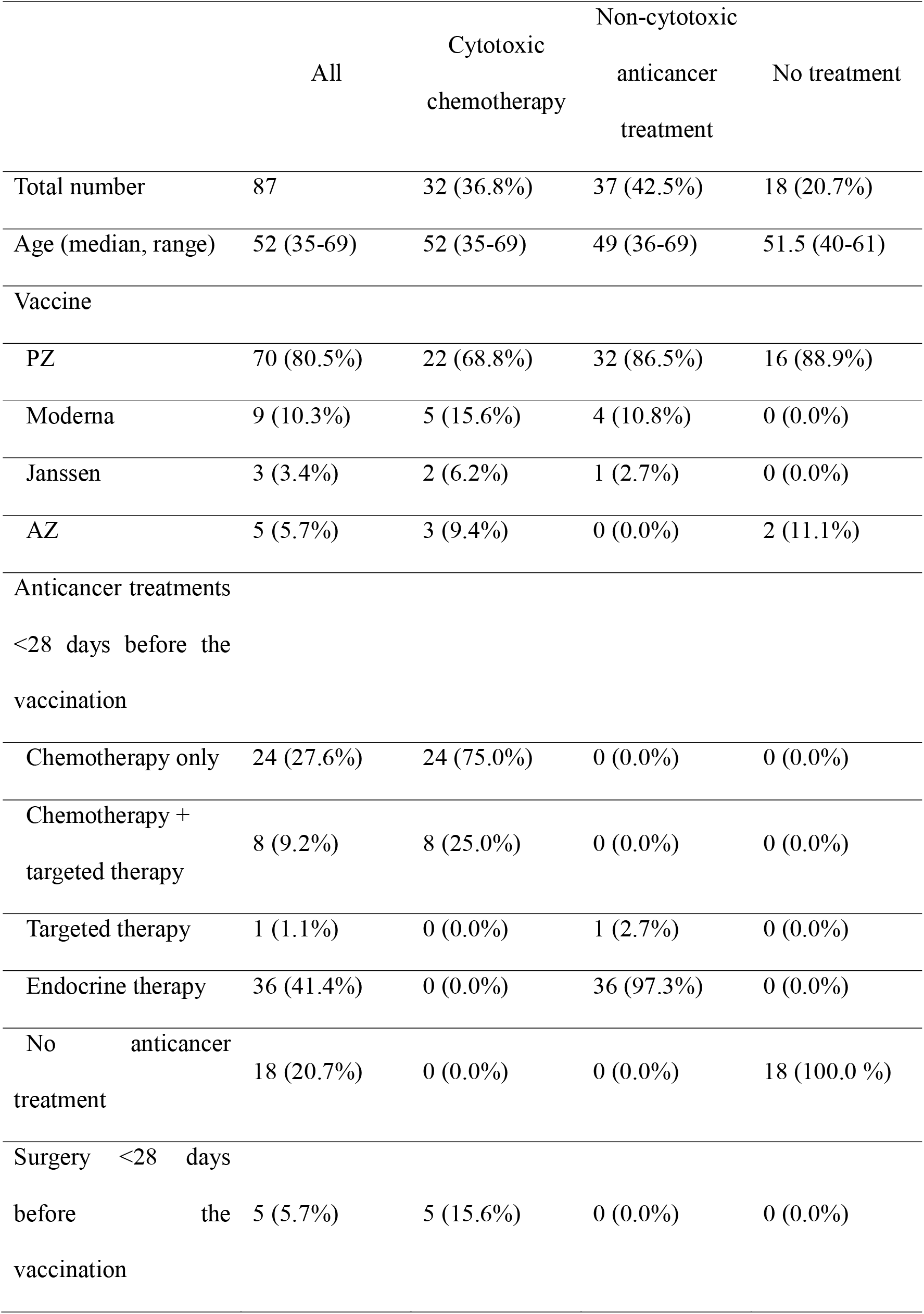
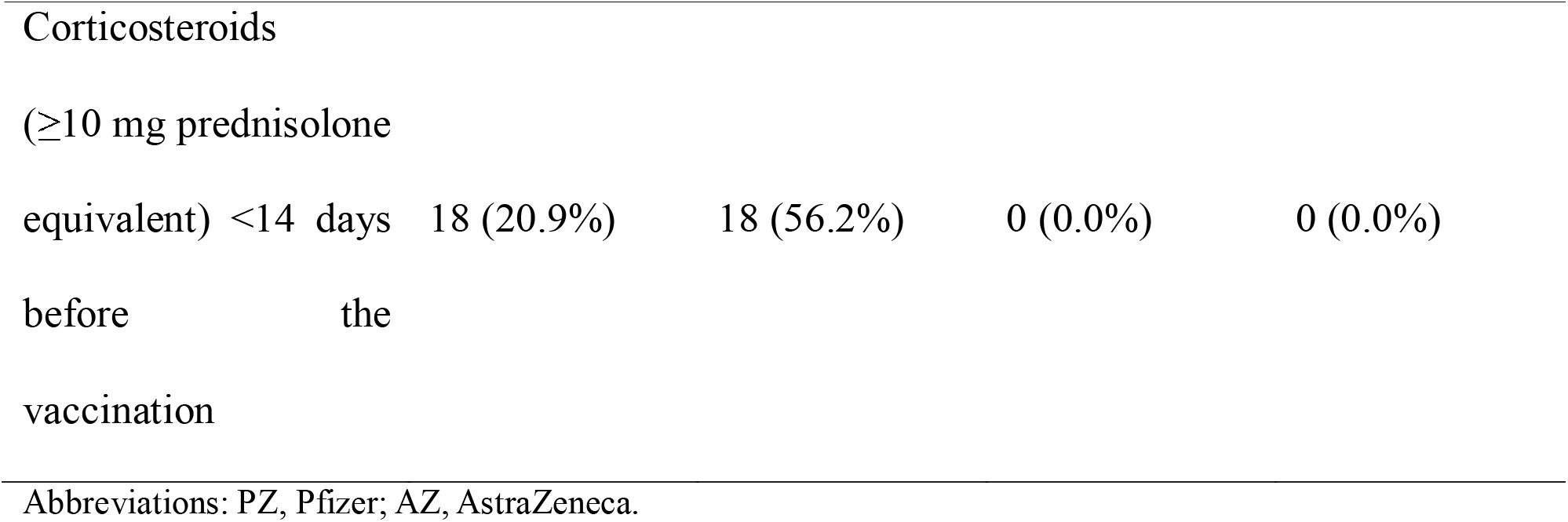
Patients’ characteristics

**Table 2.**
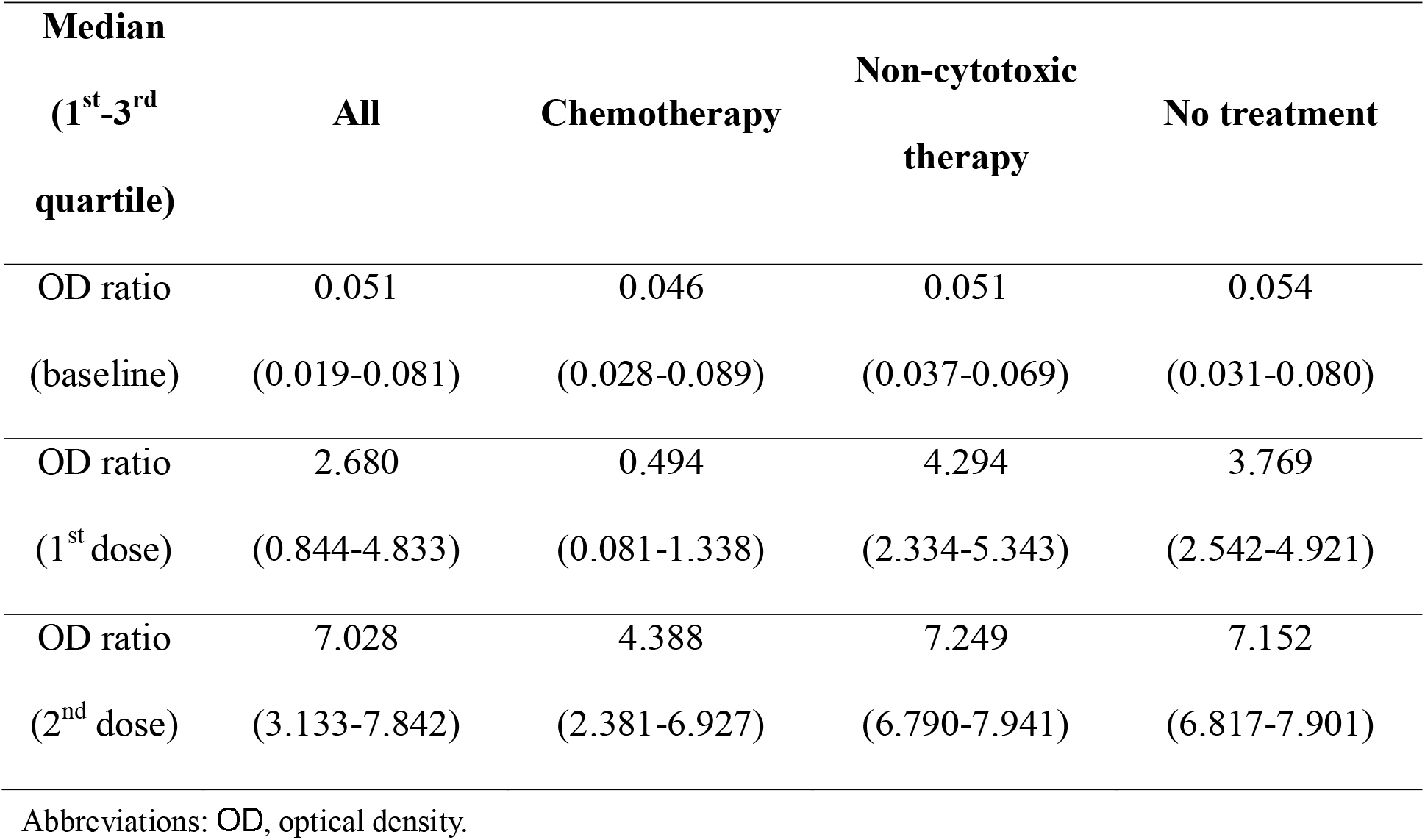
Antibody titers with Anti-Cancer treatments

### Vaccine immunogenicity with corticosteroid use

Corticosteroid was used within 14 days before the vaccination in 20.9% (18 of 87) of the patients, all of whom were in the chemotherapy group. The association between corticosteroid use and antibody titer was only assessed in chemotherapy group to control the effect of chemotherapy to immunogenicity. There was no significant difference in the antibody titers between the patients who received corticosteroid in concurrence with chemotherapy and those who only received chemotherapy at the time of vaccination (Figure 4).

**Figure 4.**
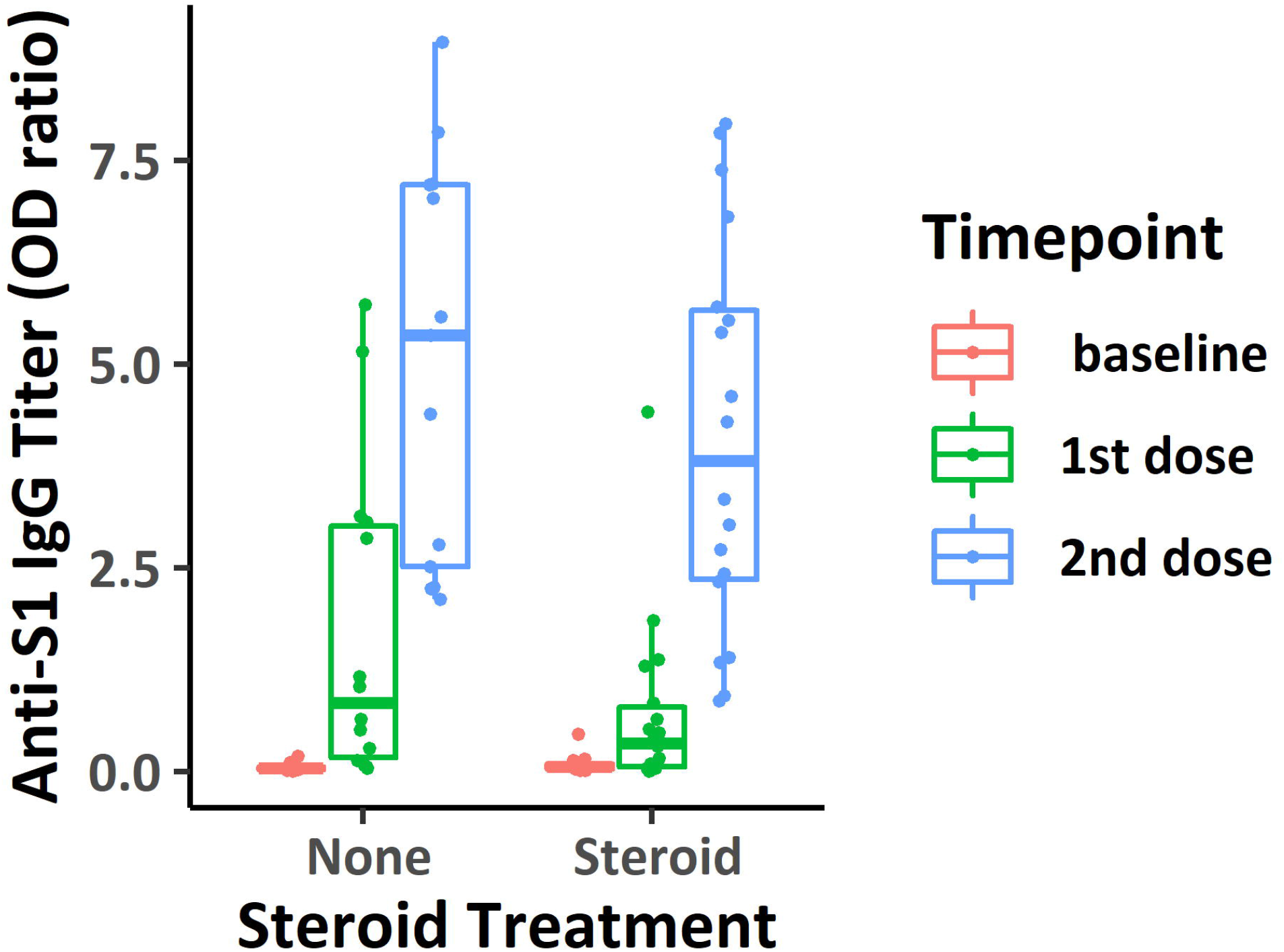
Antibody titers with steroid treatments in chemotherapy group: Comparison of SARS-CoV-2 anti-S IgG antibody titers after each dose of vaccinations with and without corticosteroid treatments in cytotoxic chemotherapy group

## Discussion

In this prospective study, standard regimen of COVID-19 vaccination generated sufficient amount of IgG antibody titers against SARS-CoV-2 virus in most of the early breast cancer patients who had adjuvant anti-cancer treatments. The rate of seropositivity was numerically comparable to that of the previous studies performed in patients with cancer ^13,14,18,19^. It is well known that the anti-spike antibody titer is correlated with the protection from COVID-19 ^20-23^. The emergence of neutralizing antibodies at early onset was associated with the shorter time for negative conversion of swab culture, while the lack of neutralizing antibody was associated with a fatal outcome in symptomatic COVID-19 patients ^20^. Known as a good surrogate marker of neutralizing antibody titers ^20,23^, the anti-spike antibody levels measured with EUROIMMUN ELISA kit also reported strong positive correlation with the neutralizing antibody titers assessed by a microneutralization assay ^21^. All in all, patients with breast cancer under concurrent adjuvant cytotoxic chemotherapy, when compared with those under molecular targeted therapy, endocrine therapy, or no anti-cancer treatments, were capable of acquiring adequate protections against COVID-19 infection after vaccinations, irrespective of age and types of vaccines.

Cytotoxic chemotherapy, however, was significantly associated with lower IgG antibody titers after both first and second dose of vaccinations despite the equivalent seroconversion rate. About 7.8-fold difference was observed between the cytotoxic chemotherapy group and non-cytotoxic treatment group after the first dose of vaccine, which decreased to 1.5-folds after the second dose. A cohort study reported that the anti-spike IgG titers of patients with solid cancer was 3.7-folds lower than those of healthy relatives after the second dose of Pfizer vaccination ^13^. Moreover, two of the patients were under adjuvant cytotoxic chemotherapy before PZ vaccine showed borderline range of anti-S1 IgG levels. In PZ vaccination trial, the median IgG titer was statistically significantly lower in the patients under active cancer treatments with healthy controls ^13^. Booster dose of vaccinations may be considered in the cancer patients under active cytotoxic treatments for a robust protection against SARS-CoV-2.

The only patient who was seronegative had received one dose of Janssen while on endocrinal therapy, and had completed cytotoxic chemotherapy 47 days before vaccination. Meanwhile, two other patients who had received Janssen vaccine experienced sufficient seroconversion after the first dose between cycles of cytotoxic chemotherapies. One possible speculation would be their ages. The seronegative patient was over 65 years old while other two seropositive patients were in their 40s and 50s. In the phase I/II trial of Janssen vaccine, the participants of 60 years or older had lower incidence of seroconversion ^24^. In the subgroup analysis of ENSEMBLE trial, lower efficacy of preventing the severe-critical COVID-19 was observed among participants of 60 years or older with comorbidities ^6^. Individual medical circumstances should be taken into account in choosing the type of vaccines, and booster doses should be considered for vulnerable populations.

The non-cytotoxic treatment group treated with endocrinal therapy and monoclonal anti-HER-2 therapy showed similar seroconversion rates and anti-S1 antibody titers when compared to control group. Previous study reported higher rates of post-vaccination seroconversion in patients on endocrine therapy including ovarian function suppression and aromatase inhibitors compared with the other groups ^18^. Only scarce data was available regarding the impact of the trastuzumab to the efficacy of the vaccines. One study about influenza vaccination reported that breast cancer patients receiving adjuvant trastuzumab had sufficient increase of immunogenicity and equivalent serological protection rate as healthy controls ^25^.

It had been widely believed that the use of systemic corticosteroid may affect T and B cell functions. For example, patients with sustained high-dose corticosteroids were associated with poor pneumococcal polysaccharide vaccine response and more infections ^26^. In our study, corticosteroids over 10mg of prednisolone or equivalent dose within 14 days of vaccination apparently seemed to diminish the amount of antibody titers. But corticosteroids were administered as the premedication for taxanes, and none of the control group used corticosteroids. When the factor of cytotoxic chemotherapy was adjusted, there was no difference in the antibody titers with corticosteroid treatments. A recent study also reported that the short term use of corticosteroids significantly relieved side effects without hindering immunogenicity after the first dose of AZ vaccine ^27^. In our study, adjunctive corticosteroids were used shortly, and thus minimally impeded the immunologic responses driven by vaccines.

Our data is limited by the relatively small number of patients, especially those who had adenovirus vector vaccines. Unfortunately, only one patient was treated with trastuzumab that the analysis of the effect of molecular target therapy on immunogenicity of vaccines was unavailable. Immune checkpoints inhibitors, which are not yet approved as adjuvant treatment for breast cancer, were not included in the study. However, this study was successful in recruiting a homogeneous population of the early breast cancer patients undergoing adjuvant anti-cancer treatments. We also evaluated the effects of corticosteroid use to the seroconversion, which is commonly used in adjunct to cytotoxic chemotherapy. Another pitfall was the possibility of the seroconversion by asymptomatic COVID-19 infections, because the regular COVID-19 PCR screenings were not done. T cell responses, known as important in immunity against COVID-19, were also not evaluated. Further studies with more patient data would strengthen our knowledge in understanding the immunologic responses after vaccination in the patients with cancer.

## Conclusion

Patients with early breast cancer undergoing adjuvant anti-cancer therapy showed adequate serologic responses against SARS-CoV-2 virus after vaccination without serious adverse events. Selection of the COVID-19 vaccines should be personalized, taking the patient’s medical circumstances into account. Booster doses of vaccination could be considered for the vulnerable populations.

## Data Availability

All data produced in the present study are available upon reasonable request to the authors.

## Supplementary Figures & Tables

Figure S1. Antibody titers with vaccinations in all patients: SARS-CoV-2 anti-S IgG titers after each dose of vaccinations in all early breast cancer patients

Figure S2. Antibody titers by vaccine types: SARS-CoV-2 anti-S IgG titers after each dose of Adenoviral vaccines and mRNA vaccines

Figure S3. Antibody titers with steroid use: SARS-CoV-2 anti-S IgG titers in adjunct with steroid use of more than or equivalent to prednisolone 10mg within 14 days of each dose of vaccinations

Table S1. Seroprevalence of early breast cancer patients

Table S2. Characteristics of Patients who Failed Seroconversion

Table S3. Univariable logistic regression analysis for seropositivity

Table S4. Univariable Linear Regression Analysis for OD values

Table S5. Multiple Regression Analysis for OD values

## Data availability

The datasets generated during and/or analyzed during the current study are available from the corresponding author on reasonable request.

## Acknowledgments

We thank Areum Jo for technical support.

