## Supplementary figures and images for "Evaluation of Seropositivity after Standard Doses of Vaccination against SARS-CoV-2 in Early Breast Cancer Patients Receiving Adjuvant Treatment"

### Supplementary Figure 1

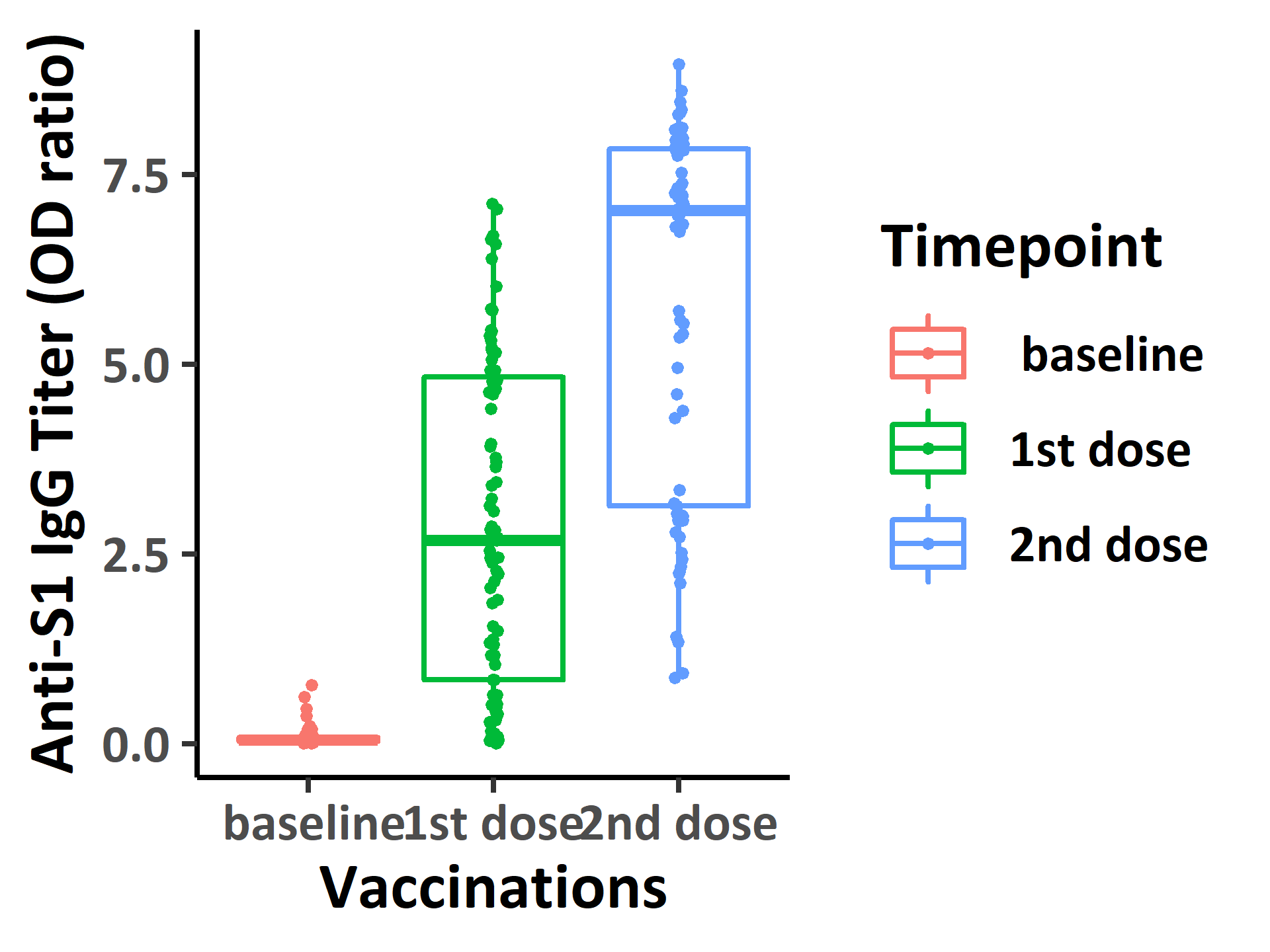

### Supplementary Figure 2

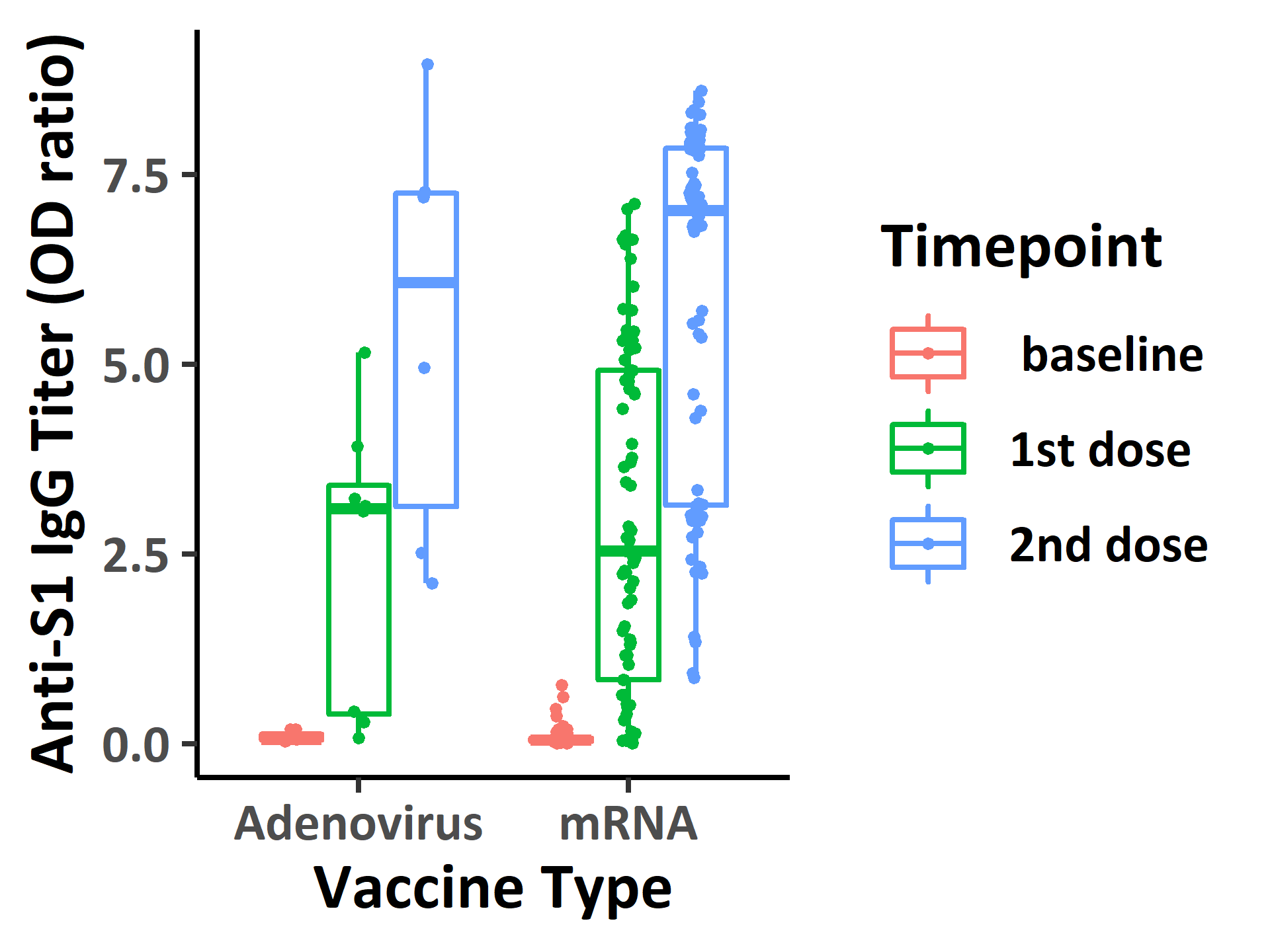

### Supplementary Figure 3

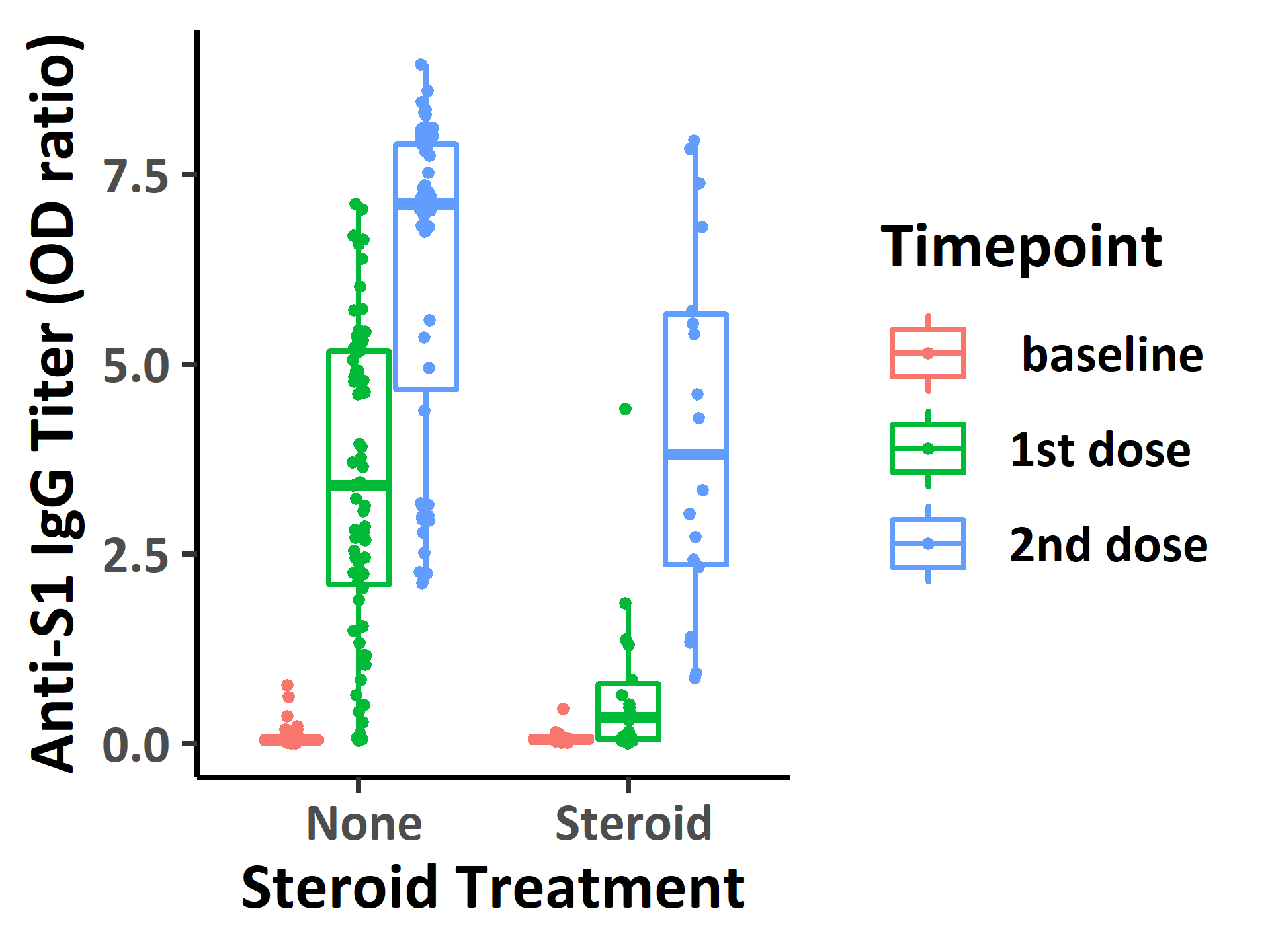
