## Supplementary Table 1 for "Evaluation of Seropositivity after Standard Doses of Vaccination against SARS-CoV-2 in Early Breast Cancer Patients Receiving Adjuvant Treatment"

Table S1. Seroprevalence of early breast cancer patients

| **Serology** | **All** | **Chemotherapy** | **Non-cytotoxic therapy** | **No treatment** |
| --- | --- | --- | --- | --- |
| **After 1^st^ dose** |  |  |  |  |
| Positive | 61 (70.1%) | 10 (31.2%) | 34 (91.9%) | 17 (94.4%) |
| Borderline | 3 (3.4%) | 2 (6.2%) | 1 (2.7%) | 0 (0.0%) |
| Negative | 21 (24.1%) | 20 (62.5%) | 1 (2.7%) | 0 (0.0%) |
| NA | 2 (2.3%) | 0 (0.0%) | 1 (2.7%) | 1 (5.6%) |
| **After 2^nd^ dose** |  |  |  |  |
| Positive | 83 (95.4%) | 29 (90.6%) | 36 (97.3%) | 18 (100%) |
| Borderline | 2 (2.3%) | 2 (6.2%) | 0 (0.0%) | 0 (0.0%) |
| N/A | 2 (2.3%) | 1 (3.1%) | 1 (2.7%) | 0 (0.0%) |
| **Overall** |  |  |  |  |
| Positive | 84 (96.6%) | 30 (93.8%) | 36 (97.3%) | 18 (100%) |
| Borderline | 2 (2.3%) | 2 (6.2%) | 0 (0.0%) | 0 (0.0%) |
| Negative | 1 (1.1%) | 0 | 1 (2.7%) | 0 (0.0%) |

Abbreviations: N/A, not available.
