## Supplementary Table 2 for "Evaluation of Seropositivity after Standard Doses of Vaccination against SARS-CoV-2 in Early Breast Cancer Patients Receiving Adjuvant Treatment"

Table S2. Characteristics of patients who failed seroconversion

| Patient | Vaccine | Anti-cancer treatment | Vaccine interval (days) | Steroid <14 days  before the vaccination | NSAIDs <14 days  before the vaccination | Serology |
| --- | --- | --- | --- | --- | --- | --- |
| 1 | Janssen | Femara | N/A | No | N/A | Negative |
| 2 | PZ | Docetaxel after AC | 21 | Yes | No | Borderline |
| 3 | PZ | Docetaxel after AC | 32 | Yes | Yes | Borderline |

Abbreviations: PZ, Pfizer.
