## Supplementary Table 3 for "Evaluation of Seropositivity after Standard Doses of Vaccination against SARS-CoV-2 in Early Breast Cancer Patients Receiving Adjuvant Treatment"

Table S3. Univariable logistic regression analysis for seropositivity

| Variable | p-value | OR (95% Confidence interval) |
| --- | --- | --- |
| Age | 0.661 | 0.965 (0.821-1.132) |
| Vaccine type  (mRNA or adenovirus) | 0.185 | 0.182 (0.015-4.195) |
| Chemotherapy | 0.304 | 3.600 (0.332-79.372) |
