## Supplementary Table 4 for "Evaluation of Seropositivity after Standard Doses of Vaccination against SARS-CoV-2 in Early Breast Cancer Patients Receiving Adjuvant Treatment"

Table S4. Univariable Linear Regression Analysis for OD values

| Variable | B | SE | β | t-value | p |
| --- | --- | --- | --- | --- | --- |
| Age | 0.086 | 0.0.034 | 0.265 | 2.506 | 0.014 |
| Vaccine type  (mRNA or adenovirus) | -0.354 | 0.997 | -0.039 | -0.355 | 0.724 |
| Chemotherapy | 2.103 | 0.478 | 0.435 | 4.400 | <0.001 |
