## Supplementary Table 5 for "Evaluation of Seropositivity after Standard Doses of Vaccination against SARS-CoV-2 in Early Breast Cancer Patients Receiving Adjuvant Treatment"

Table S5. Multiple Regression Analysis for OD values

| Variable | B | SE | Β | t-value | p |
| --- | --- | --- | --- | --- | --- |
| Age | 0.088 | 0.031 | 0.270 | 2.851 | 0.006 |
| Chemotherapy | 2.057 | 0.435 | 0.438 | 4.728 | <0.001 |
